# Acceptability and appropriateness of remote patient monitoring and self-administered pulse oximetry among COVID-19 patients in Honduras: a mixed methods study

**DOI:** 10.1101/2025.06.22.25330090

**Authors:** Kathryn W. Roberts, Berta Alvarez, Omar Diaz, Michael de St. Aubin, Salome Garnier, Saul Cruz, Lorenzo Pavon, Shiony Midence, Angela Ochoa, Homer Mejia Santos, Jonatan Ochoa, Sogeiry Solis, Rachel See, Devan Dumas, Margaret Baldwin, Ligia Paina, C. Daniel Schnorr, Alcides Martinez, Avi J Hakim, Eric Nilles

**Affiliations:** Harvard Humanitarian Initiative, 14 Story Street, 2nd Floor, Cambridge, MA 02138; Mass General Brigham Hospital, 75 Francis Street, Boston, MA 02115; Johns Hopkins Bloomberg School of Public Health, Baltimore, MD 21205; Secretaria de Salud de Honduras. Dirección General de Redes Integradas de Servicios de Salud; Secretaria de Salud de Honduras. Unidad de Vigilancia de la Salud; Centers for Disease Control and Prevention, COVID-19 International Task Force; Harvard University Medical School, 25 Shattuck St, Boston, MA 02115

**Keywords:** COVID-19, telehealth, remote patient monitoring, Honduras, pulse oximeters, acceptability, appropriateness, mixed-methods, implementation science

## Abstract

**Introduction:** During the COVID-19 pandemic, the Honduras Secretariat of Health (SESAL) and partners conducted a randomized trial to assess the impact of remote patient monitoring with versus without self-administered pulse oximetry in high-risk populations in urban Honduras. Acceptability and appropriateness were examined to inform future adaptation of the intervention to other cultural and epidemiological contexts where remote monitoring and home-based technology use can reduce barriers to healthcare access.

**Methods:** This mixed methods study included trial participants, staff, and SESAL staff working in COVID-19 triage centers. Data sources include trial data, computer-aided self-interviews, and in-depth interviews. The trial was implemented between March 2022 and January 2023. Quantitative and qualitative integration occurred during study design, analysis, interpretation, and reporting, and a joint display enables analysis and interpretation.

**Results:** 1,767 study participants completed a discharge questionnaire. Thirty-four healthcare providers completed a computer-aided self-interview, and 16 of these additionally participated in an in-depth qualitative interview.

Participants and providers demonstrated strong understanding of interventions and expressed positive affective attitudes, although SESAL staff attitudes were somewhat less positive. Trial participants were satisfied with the care they received and 94.9% reported a willingness to participate in the future. Some study and SESAL staff expressed concern over participant comprehension and self-administration of pulse oximeters, which led some to report reluctance to enroll patients in the future. In contrast, participants reported successfully using and understanding pulse oximeter readings. Healthcare providers were confident in their ability to implement the intervention in a clinical environment, but a few expressed concerns about its appropriateness, when asked to consider its implementation versus competing health priorities.

**Conclusion:** Overall, study participants and healthcare providers found remote monitoring and self-administered pulse oximetry acceptable ways to monitor for deterioration during the acute phase of a COVID-19 infection. Findings showed that an intervention can be appropriate from a participant perspective but that appropriateness may be less clearcut from a provider perspective due to competing priorities, despite the interventions being highly acceptable. The approach shows promise for adaptation in other settings experiencing health emergencies where technology penetration is high and healthcare availability does not align with demand, but appropriateness considerations should be explored prior to implementation.

**What is already known about this topic:** (Summarize the state of scientific knowledge on this subject before you did your study and why this study needed to be done)

Uptake of remote patient monitoring and telehealth in Latin America accelerated during the COVID-19 pandemic, however, most descriptions of these processes and their outcomes do not include acceptability and appropriateness assessments. The acceptability and appropriateness of self-administered pulse oximetry for COVID-19 patient monitoring in LMIC has been questioned given limited local resources, lack of quality-assured devices, and low reading and health literacy, which could lead to improper use or inability to interpret results.

**What this study adds:** (summarize what we know now because of this study that we did not know before)

The high levels of acceptability among both participants and healthcare providers demonstrate that remote monitoring, with or without self-administered pulse oximetry, can be acceptable in LMIC if interventions are adapted to the local context using feedback from target population, and education and ongoing support are provided.

**How this study might affect research, practice, or policy:** (summarize implications of the study)

Assessing the acceptability and appropriateness of these interventions facilitates future adaptation and application, promoting remote healthcare access for patients with mild illnesses in contexts where health systems are overburdened and telephone-based communication is prevalent, whether during health emergencies or to monitor patients with acute or chronic conditions at home.

## 1 Introduction

Epidemics frequently overwhelm healthcare systems, exceeding capacity to deliver safe and appropriate care.(1,2) During the COVID-19 pandemic, hospitals in Honduras and globally were stretched beyond capacity by case surges.(3) In response, the World Health Organization (WHO) released guidelines describing recommended approaches for home-based management of stable patients at high risk for severe COVID-19 when hospitalization was unavailable.(4) The Honduras Secretariat of Health (SESAL) adapted those guidelines for national use and conducted a randomized trial to assess the impact of remote patient monitoring, with versus without self-administered pulse oximetry, in COVID-19 patients with mild disease at high risk for adverse outcomes.(5,6) This study analyzes data from that trial and additional interviews with healthcare providers to explore intervention acceptability and appropriateness.

In Latin America, the adoption of telehealth accelerated during the COVID-19 pandemic, however, most descriptions do not include implementation outcomes.(7–9) A systematic review of telehealth adoption during the COVID-19 pandemic found that it offered clinical, organizational, technical, and social opportunities like improving safety and care effectiveness, reducing hospitalization, maintaining continuity of care, and improving patient satisfaction. An examination of telehealth’s possible benefits and challenges in Honduras during COVID-19 emphasized the importance of accessibility, ethicality, and confidentiality.(10) Challenges include privacy and confidentiality concerns, lack of physical exams, difficulty connecting to communications networks, and gaps in technology skills.(11) The acceptability and appropriateness of self-administered pulse oximetry for COVID-19 patient monitoring in LMIC has been questioned given limited local health system and patient resources, lack of quality-assured devices, and low reading and health literacy, which could lead to incorrect use or results interpretation.(12) However, analyses of self-administered pulse oximetry for COVID-19 and other respiratory diseases, including the parent trial to this study, have demonstrated correct use and accurate interpretation, given appropriate education and consistent support.(13)

The field of implementation science emerged to study methods to promote and evaluate the systematic uptake of research findings and evidence-based practices into healthcare.(14,15) Acceptability and appropriateness are central constructs, influencing the adoption and sustainability of new interventions.(16) Acceptability of healthcare interventions can be examined through Sekhon et al.’s theoretical framework, which is comprised of constructs that reflect “the extent to which people delivering or receiving a healthcare intervention consider it to be [acceptable], based on anticipated or experiential cognitive and emotional responses to the intervention”.(17) Acceptability refers to the perception among patients, providers, and policymakers that an intervention is agreeable or satisfactory.(17) Appropriateness considers the perceived fit or relevance of an intervention for a specific setting, population, or problem.(16) Understanding the acceptability and appropriateness of remote patient monitoring and self-administered pulse oximetry is crucial to promote future uptake and adoption. This approach could be employed during health emergencies across cultural or epidemiological contexts to monitor patients recovering from acute illness or acute stages of chronic conditions.

## 2 Methods

### 2.1 Study design and data sources

This mixed methods study uses data collected during a pulse oximetry randomized trial and questionnaire and interview data collected after study completion. An exploratory sequential mixed methods design was chosen to provide depth and breadth to the investigation of these complex topics. Acceptability was assessed using domains described by Sekhon et al.(17,18) **(Figure 1)**

**Figure 1:**
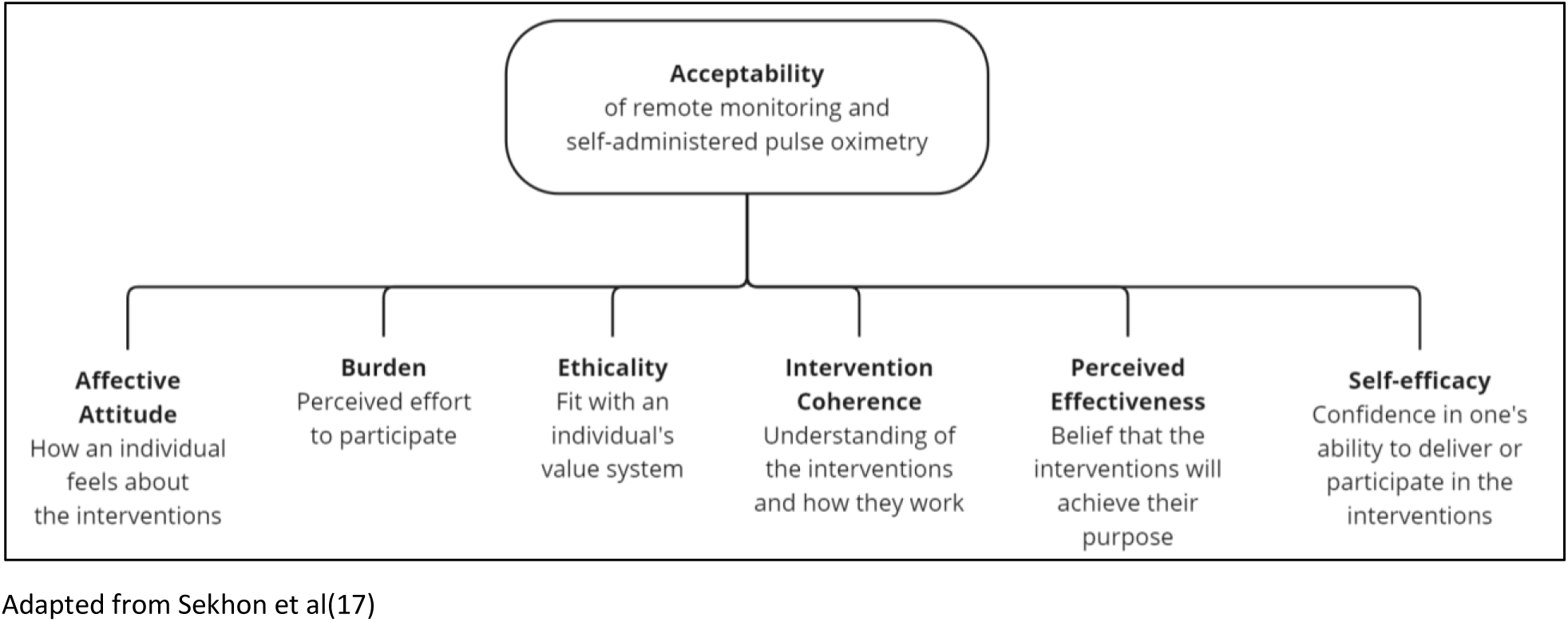
Constructs of acceptability examined about remote patient monitoring and self-administered pulse oximetry.

The integration of quantitative and qualitative methods occurred during study design, analysis, interpretation, and reporting, as described by Fetters et al.(19) Trial participants responded to quantitative questions, allowing researchers to draw conclusions about the study population, while open-ended questions permitted participants to express opinions. Healthcare providers responded to a preliminary, anonymous, computer-aided self-interview (CASI) including quantitative and open-ended questions exploring study perceptions, given limited information about their perspectives. (16,17) CASI responses were used to develop an in-depth interview questionnaire, which enabled examination of rationale for perspectives expressed in the CASI. The results from participant and healthcare providers were integrated through a joint display to compare experiences and perceptions of acceptability and appropriateness. **(Figure 2)**

**Figure 2:**
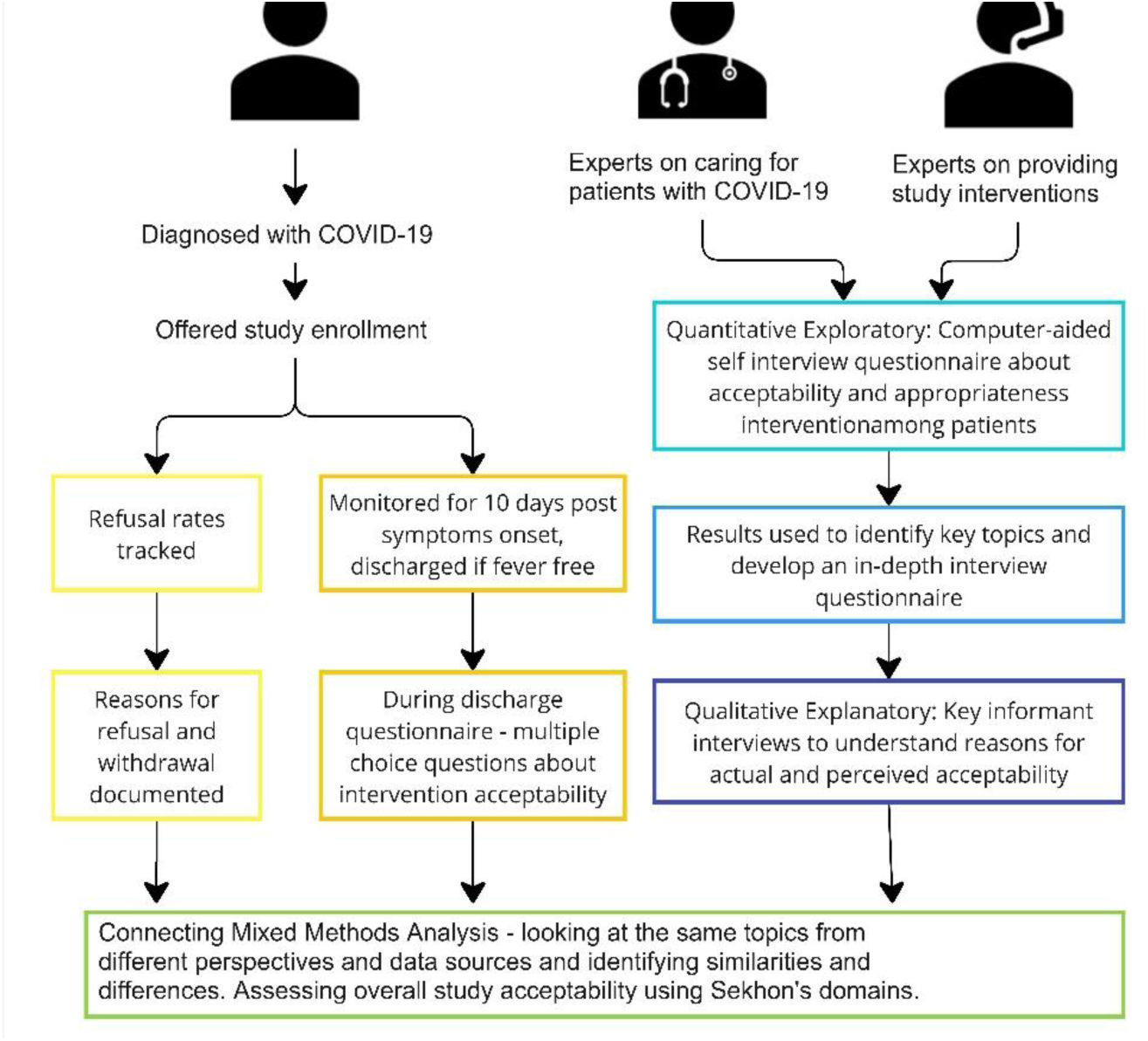
**Acceptability and appropriateness sub-study methods**.

### 2.2 Randomized trial

Between March 30, 2022 and January 24, 2023, 1,821 participants were enrolled in a pragmatic, block-randomized trial in Honduras’ capital city Tegucigalpa and the adjacent city of Comayagüela, to assess the impact of remote monitoring with versus without self-administered pulse oximetry on COVID-19 patients at high risk for adverse outcomes. Participants were enrolled at five COVID-19 triage centers managed by SESAL and the Social Security Hospital Institute, the only sites in intervention cities where public sector COVID-19 testing was available at trial commencement. Eligibility criteria included COVID-19 symptoms, a positive SARS-CoV-2 rapid antigen test (Roche Diagnostics, Switzerland), living or working in Tegucigalpa or Comayagüela, access to a functional telephone, not requiring hospitalization at the time of evaluation as determined by the treating SESAL physician, and aged 60 years or older or 45-59 years of age with one or risk factors for severe COVID-19. (20) **(S1)** Participants were block randomized to receive remote monitoring with or without self-administered pulse oximetry, in which case they received an FDA-approved pulse oximeter procured in the U.S., costing ∼$35.Thereafter, study nurses called participants daily to assess warning signs, including Sp02 ≤ 94% for those with pulse oximeters. Participants reporting warning signs were immediately referred to a COVID-19 triage center for in-person evaluation. Participants were disenrolled ten days post symptom onset unless the participant was admitted to a hospital for > 24 hours or reported subjective or measured fever (≥38.0° C) during the prior 24 hours, in which case daily monitoring continued until they were fever-free for 24 hours.

### 2.3 Study procedures

All patients who met inclusion criteria but declined to participate or withdrew after enrollment were invited to complete a brief questionnaire. Trial participants that completed remote monitoring were administered a disenrollment questionnaire that included acceptability.

CASI eligibility included SESAL physicians and physician administrators familiar with the trial and working at COVID-19 triage centers when data were collected, and all trial physicians and nurses. CASI data were collected in November and December 2022. The electronic questionnaire was developed in Kobo Toolbox and distributed to SESAL staff in-person on tablets and to study staff through What’s App.(21). Participants could remain anonymous or provide contact details if willing to be interviewed.

In-depth interviews participants were selected using purposeful, segmented, maximum variation sampling from the pool of CASI participants willing to participate. Participants were segmented by employer, role, and gender, and 16 selected to participate, all of whom accepted. The Spanish-language interviews were framed by Sekhon et al’s acceptability constructs, inviting providers to share personal experiences with and perceptions of participants’ engagement with the interventions, explored rationale for beliefs identified through the CASI, and defined and discussed intervention appropriateness. Saturation was defined as encountering repeated experiences and perceptions, such that researchers judged additional data collection unlikely to surface disparate perspectives. Senior study staff conducted in-depth interviews in-person or by phone after trial enrollment was complete. Interviews took place in a private space where they were audio-recorded, and a second staff member simultaneously took notes. Note-takers created verbatim transcripts; transcript accuracy was verified by the interviewer.

### 2.4 Informed consent and ethical approval

Study participants provided written or electronic informed consent prior to participation. Eligible trial participants that declined enrollment provided verbal consent prior to completing the refusal questionnaire. Study and SESAL staff provided electronic written consent. In-depth interview participants provided written consent when interviewed in person and verbal consent when interviewed over the phone, with the interviewers providing signatures affirming consent was obtained. This study was approved by the Biomedical Research Ethics Committee of the Faculty of Medical Sciences, National Autonomous University of Honduras, Tegucigalpa (Approval No. 00003070), and the Mass General Brigham Human Research Committee, Boston, USA (Approval No. 2021P001143), and secondary data analysis was approved by the Johns Hopkins Bloomberg School of Public Health (Approval No. 24586). The protocol was reviewed by the CDC GHC ADS and determined to be research but CDC was not engaged.

### 2.5 Data analysis

Quantitative analysis of trial participation refusal and withdrawal rates were summarized and logistic regression used to analyze demographic differences compared to participants. Quantitative trial disenrollment data was analyzed using logistic regression with difference assessed by sociodemographic variables, intervention assignment, and intervention dose received. Quantitative CASI data were summarized and Fisher’s exact test was used to compare differences in responses to each question by employer, role, gender, and age. Responses to open-ended questions were thematically categorized and summarized separately from other qualitative data collected. Quantitative data were analyzed in STATA/17.0, 2021 and R 4.2.2, 2022.(22,23).

In-depth interviews were analyzed using deductive framework analysis grounded in Sekhon et al’s work with space for inductive code generation based on discussion between the two coders and framework functionality in relation to the data. A codebook was developed to ensure transparency and coding alignment. Coders used regular discussion and collaborative memoing, where qualitative researchers record thoughts, insights, and initial interpretations in shared documents, to ensure reflexivity.(24) Each coder reviewed all transcripts for framework and code development. Next, each coded two transcripts, reviewed the other’s work, discussed disparities in code application, and updated code definitions. Then, each led coding on half the remaining transcripts, which by the coder then reviewed. Qualitative data were organized in Dedoose and Microsoft Excel.(25,26) All data were collected in Spanish and all textual data analyzed in Spanish, with results translated for this manuscript by K. Roberts.

Following analysis of each data source, results were visualized in a joint display.(27) The data types are separated along the x-axis and organized thematically by Sekhon et al’s domains of healthcare acceptability and appropriateness on the y-axis. Within the joint display, qualitative interview results are summarized for brevity. Following joint display population, data were examined by domain to understand experiences and perceptions of intervention acceptability, rather than to triangulate findings.

## 3 Results

Among 1,860 COVID-19 positive patients screened for enrollment, 1,841 met study criteria, of which 19 (1.0%) declined enrollment. 1,821 people were enrolled and one was enrolled twice. The profile of those that declined enrollment was similar to those enrolled. **(Table S2)** 1,768 (97.1%) study participants completed the intervention and were discharged per protocol, 48 were lost to follow-up (2.6%) and five (0.2%) withdrew before reaching trial endpoints. 1,767 (97.0%) of those discharged per protocol completed a discharge questionnaire. **(Figure 3)**

**Figure 3:**
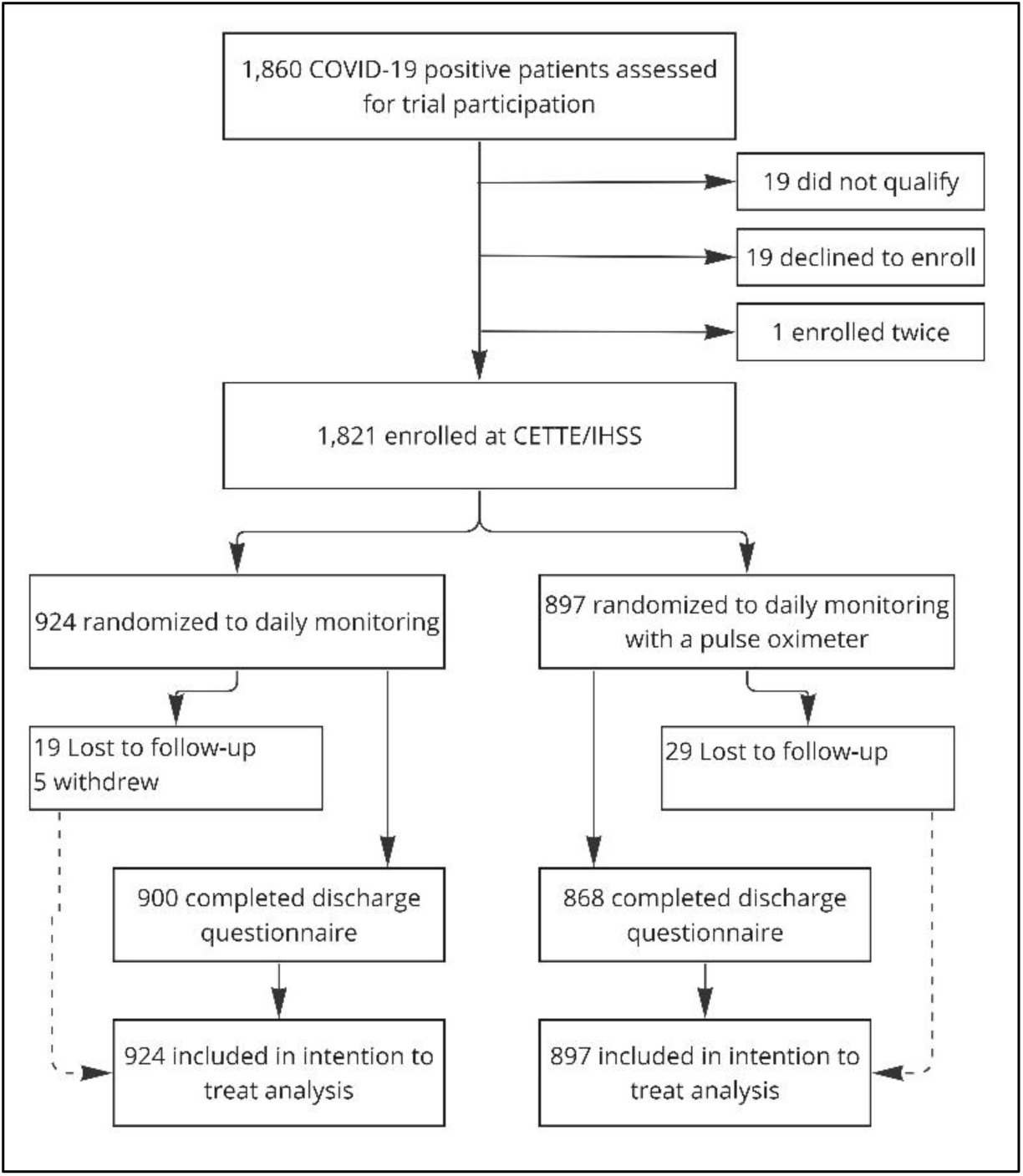
Trial profile.

Thirty-four people completed the CASI questionnaire, 19 SESAL staff (three physician administrators and 16 physicians) and 15 study staff (eight physicians and seven nurses) **(Table S3)**. Thirty-two of 34 CASI participants volunteered for an in-depth interview, of which eight SESAL and eight study staff were selected and interviewed. **(Table S4)** Following interview coding, coders excluded one interview due to lack of respondent familiarity with the intervention. Given small sample size, age and gender are not reported with quotes for respondent privacy.

**Table 1:** Joint display of trial participant, study staff, and SESAL employee perceptions of acceptability and appropriateness of remote patient monitoring and self-administered pulse oximeter use.

### Intervention Coherence

Intervention coherence describes the extent to which participants and providers understood how the intervention was intended to function. Participants were asked the meaning of pulse oximeter results and healthcare providers were asked to describe intervention objectives during in-depth interviews. SESAL and study staff that participated in in-depth interview understood intervention objectives, explaining the intention to educate patients about COVID-19 warning signs, provide support during recovery through warning signs identification and pulse oximeter use, followed by referral for evaluation. Respondents highlighted the relationship between study participation and improved health outcomes. SESAL and study staff frequently believed the intervention was designed to support mental health, although it was not, in fact, an intervention objective. One SESAL doctor believed that the study aimed to reduce unnecessary returns for care, and therefore overall patient volume at COVID-19 triage centers.

Trial participant understanding of pulse oximeters was assessed during disenrollment. Among those in the pulse oximetry arm that completed the enrollment period, 99.4% (n=841) reported that pulse oximeters measured blood oxygen levels. Among respondents who gave different answers, three reported the pulse oximeter was broken, one that it was slow, and one said the device told them that “everything was okay”. No respondents believed the device measured something other than blood oxygen. Intervention coherence was only assessed in the pulse oximeter arm.

### Affective attitude

Affective attitude examines how participants and healthcare providers felt about the interventions. Using trial data, we assessed participant willingness to enroll, withdrawal, and satisfaction. Using CASI data, we assessed healthcare provider willingness to refer patients and family members for interventions and perceptions of participants’ affective attitude towards interventions.

Among trial-eligible patients (n=1,860) who declined to enroll (n=19), reasons relating to affective attitude included not wanting to be called (n=8), not believing follow-up was necessary (n=4), or disinterest in the interventions or research (n=2). Remaining participant reasons for declining are discussed in forthcoming sections.

Five study participants withdrew after enrolling, all in the monitoring only arm (p=0.062). Participants withdrew a mean of 4.4 days after enrollment [SD 1.9]. No participant who withdrew was referred for additional care, compared to 5.8% of participants who did not withdraw (n=102). Three participants who withdrew reported that the calls were bothersome and two did not share reasoning.

During disenrollment, 98.1% of study participants reported being satisfied with home-based care; 99.0% (n=892) in the monitoring arm and 99.5% (n=862) in the pulse oximeter arm; the difference between interventions was not significant (OR 2.17 [95% CI 0.66 – 7.09], p=0.198). Dissatisfaction related to burden and self-efficacy is discussed in forthcoming sections. All hospitalized participants who responded to the disenrollment questionnaire (8/15) were very satisfied with home-based care.

94.9% (n=1,667) of study participants reported they would participate again if re-diagnosed with COVID-19. Pulse oximeter arm participants were more willing to participate again versus those in the monitoring arm (OR 1.69 [95% CI 1.08 – 2.62], p = 0.020). Seventy-five- to 84-year-olds were least willing to participate again (OR 2.64 [95% CI 1.30 – 5.37], p=0.007), without a significant difference between interventions. The odds of being willing to participate again were 1.67 times greater among people living in Comayagüela versus Tegucigalpa ([95% CI: 1.09 – 2.57], p=0.019).

Seventy-one percent of study staff (n=10) vs 43% of SESAL staff (n=8) reported they would be somewhat or very likely to refer patients for study interventions in the future, if the choice were theirs.

**Figure 4:**
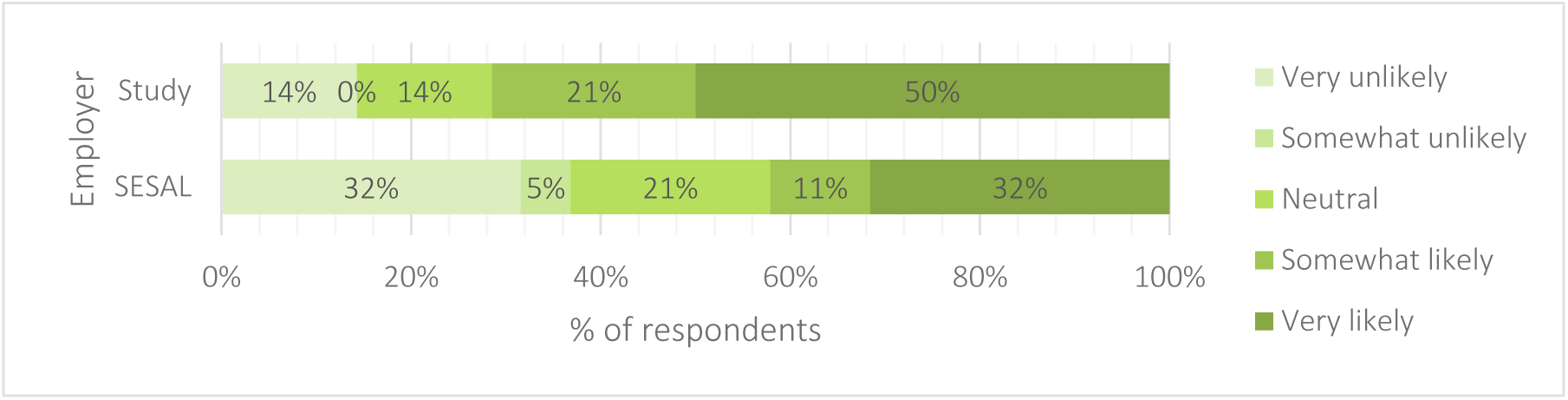
Prospective willingness to refer a qualifying patient for intervention participation.

When asked about their willingness to refer a family member with COVID-19, most SESAL (94.7%, n=18) and all study staff (100%, n=15) reporting they would be somewhat or very likely to do so.

**Figure 4:**
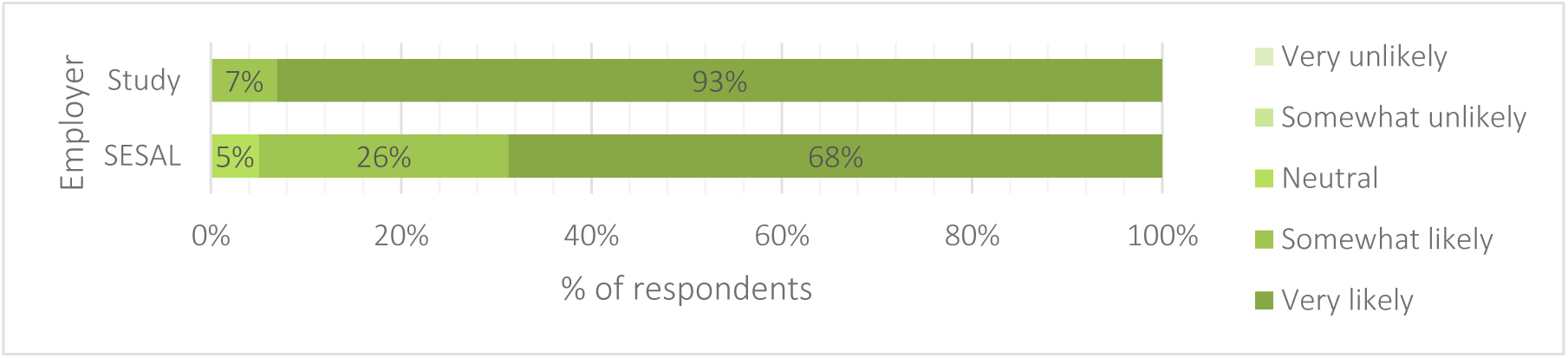
Prospective willingness to refer a qualifying family member for intervention participation.

During in-depth interviews, SESAL and study staff affective attitude towards interventions was overwhelmingly positive. Many respondents from both employers described positive participant perceptions, which influenced their own feelings. Several respondents appreciated the focus on older people, because of their risk of comorbidities, likelihood of living alone, and lower health literacy. Several people connected older people’s risk status with a need for mental and emotional support. As a SESAL doctor explained,

> *…[study participants] feel that someone is looking out for them, which is one of the most important things because you know that the emotional aspect is what practically killed many of them..*.

Four study physicians were interviewed, two shared negative affective attitudes, one believed calls were too frequent and the second believed in-person evaluation was essential.

Healthcare provider perceptions of participant experience aligned with participant reports. As one SESAL doctor described,

> *The truth is that the patients appreciate the intervention… they felt that someone was with them, that they were important…*

Providers reported that patients felt safer and more confident at home during their recovery. However, when asked to reflect on why providers were more likely to refer their own family members than patients, respondents explained that providers likely expected their family members to have more years of formal education and would receive support from the physician-relative, compared to a “usual” patient, whose family would likely be less able to provide such support.

### Perceived effectiveness

Perceived effectiveness, the belief interventions will achieve their intended purpose, was assessed by examining reasons for declining enrollment, trust in pulse oximeter results, and belief in intervention influence on quality of care.

Of the 19 potential participants who declined enrollment, one declined enrollment because they wanted a pulse oximeter, but were not assigned one. **(Table S3)** 97.4% (n=844) of participants in the pulse oximetry arm trusted device results. One of six pulse oximetry arm participants who were hospitalized reported not trusting the result, however, that participant never reported a warning sign or an Sp02 ≤94%, but went to the hospital on their own when they felt ill. 99.4% (n=861) of those with pulse oximeters believed they positively influenced quality of care, four people (0.5%) believed it was neutral, and one person (0.1%) believed the influence was negative. 98.6% (n=888) of remote patient monitoring arm participants believed the intervention had a positive influence on recovery, 0.2% (n=2) believed it had a negative influence, and 1.2% (n=11) believed it was neutral. In the CASI, 75.6% (n=25) of healthcare providers reported believing remote monitoring was an effective way to provide care. Notably, 86.7% (n=13) of study staff versus 66.7% (n=12) of SESAL staff expressed confidence in remote monitoring effectiveness. All study staff (n=15) believed the intervention had a mostly positive influence, compared to SESAL staff who believed it was mostly positive (n=13), somewhat positive (n=4), and neutral (n=2).

Study and SESAL staff emphasized that participant education enabled self-care and encouraged prevention of onward transmission. One study physician believed remote monitoring motivated participants to comply with physician recommendations. Several study and SESAL staff believed that the pulse oximeter intervention package was more effective than monitoring alone. Multiple SESAL physicians believed the interventions reduced the number of patients returning to the screening center unnecessarily. As one SESAL physician reported,

> *I think that a lot of the follow-ups that were done in triage really decreased thanks to those people who were calling them and giving them telephone follow-ups*.

Providers believed that the intervention was effective because it provided emotional support and ensured study participants did not feel alone while isolated.

### Self-efficacy

Self-efficacy is the confidence that one can perform the actions needed to carry out the intervention. Of the 19 people who declined to participate, two believed they would be unable to access a telephone. **(Table S2)** 2.3% (n=20) of trial participants in the pulse oximetry arm reported that they did not use the device. All individuals in the pulse oximetry arm that were hospitalized (n=5) reported using the device. Study participant reasons for being somewhat (n=7/1,767) or very dissatisfied (n=6/1,767) often related to self-efficacy, including difficulties accessing a phone (n=6/13), working during the call time (n=1/13), or an un-charged phone (n=1/13). The remainder did not report a reason for dissatisfaction. Among suggestions made by two or more trial participants, respondents believed this intervention should continue (n=4), remote monitoring should be used for other diseases (n=2), calls were too frequent (n=2), and calls should always originate from the same phone number (n=2).

Prior to study implementation some providers were doubtful all patients could use pulse oximeters. When interviewed after trial completion whether their perspective had shifted, one provider shared,

> *To be 100 percent honest, [in the beginning] no. Of course, the person’s schooling has a lot to do with it… [During the study] I had some people who would make little notes in their own words about how to use it or they would call a family member who would also listen to the information so they could use the oximeter well*.

All healthcare providers interviewed agreed that patients used pulse oximeters successfully and highlighted the importance of detailed instruction during enrollment, support during monitoring calls, and engaging family to ensure participants understood instructions, took Sp0_2_ readings correctly, and responded to monitoring calls.

Understanding care-seeking self-efficacy after receiving a referral was a goal of in-depth interviews, given that 46% of participants did not seek care after referral. Among respondent responses, six reasons why participants might not return were shared by at least two respondents: fear of hospitalization or dying in a hospital, fear of reinfection, previous hospital maltreatment, transportation issues (lack of funds, access, or someone to accompany them, especially among older people), reluctance to wait to see a healthcare provider at the triage center, or disbelief in warning sign seriousness.

> *Because it is not only a matter of not knowing, but also the patient’s ability to move around, because if they are people at high risk for negative outcomes who are also elderly and often cannot go out alone due to various factors, then this could also be the case*.

Descriptions of how to improve participant adherence to referrals focused on clear explanations of how warning signs connect to negative medical outcomes and reassuring the patients that they would receive timely, high-quality, attentive medical care.

Healthcare personnel expressed confidence in their self-efficacy to implement the interventions, including enrolling participants, educating about pulse oximeters, conducting remote monitoring, and providing appropriate referrals, if staffing was sufficient.

### Burden

Burden, perceptions, or experiences of effort to participate, was assessed by examining why potential participants declined enrollment or withdrew and what difficulties they experienced. Providers shared their difficulties and perceptions of patient challenges with interventions.

Of 19 potential participants who declined enrollment, two felt too ill to wait for enrollment, one declined after pulse oximeter arm assignment because they did not want to be responsible for the device. **(Table S1)** Of participants completing the discharge questionnaire, 81.9% (n=1,448) reported experiencing no difficulty participating while 18.1%(n=319) reported some difficulty. Lower educational attainment was associated with a significantly greater likelihood of reporting difficulty participating.

**Table 2:**
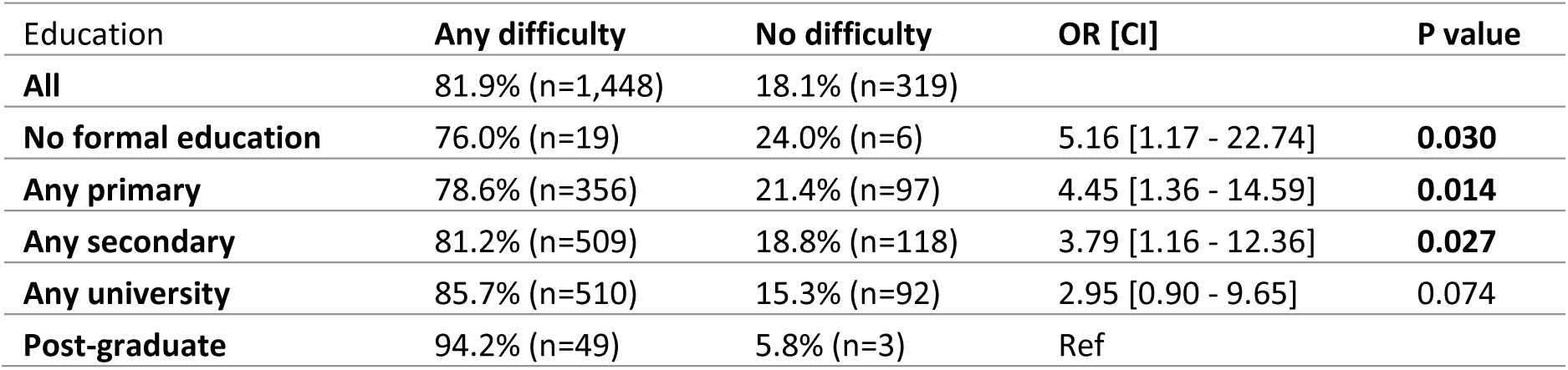
Difficulty with remote patient monitoring calls by educational attainment.

Most healthcare providers believed describing symptoms was not difficult for participants (94%, n=32). 70.6% (n=24) believed self-administered pulse oximetry was easy for participants, and 76.5% (n=26) believed it was easy for participants to read and report Sp0_2_ measurements. One SESAL physician believed waiting to be enrolled in the study was a burden for some patients. Several study nurses explained that if patients felt burdened by calls, they might complain or disenroll, which was rare.

Most (91.2%, n=31) healthcare providers did not believe enrolling participants was difficult; three SESAL respondents believed it was difficult. They believed enrollment involved some (n=12) or small effort (n=22). During in-depth interviews, SESAL physicians believed the work to implement interventions would be minimal, which some perceived as acceptable and led others to suggest increasing personnel. Study staff reported feeling overburdened by the demands of enrolling and calling patients during the height of COVID-19 waves.

### Appropriateness

Providers were asked about intervention appropriateness for the problem and context, given limited available resources and other challenges facing the population and health system. Many interview respondents shared participant approval as evidence of appropriateness, in that the interventions fit patient needs and patients appreciate the interventions. Respondents worried that while it was possible to teach participants to use pulse oximeters, they might not understand the results. One respondent expressed doubts about scalability, saying

> *…[implementing this] would imply personnel, it would imply technology and many things that SESAL does not have, there are other priorities…. Medicine, personnel…*

Another SESAL doctor believed other interventions are more urgent, like those for chronic diseases, but emphasized that monitoring high-risk COVID-19 patients is crucial because people with chronic diseases are at risk for complications.

## Discussion

This mixed methods study found that remote patient monitoring and self-administered pulse oximetry were highly acceptable to participants and healthcare providers. Both groups understood the interventions and participants were satisfied with the quality of care. Some providers expressed concerns about patient capacity to perform pulse oximetry and understand results, but participants did not share those worries. Patient educational attainment was associated with greater likelihood of reporting difficulty with an intervention, but not acceptability. While participants and providers believed monitoring patients at home was valuable, some providers wondered whether the intervention was appropriate for the context, given other urgent, non-COVID-19, health needs.

Grounding this investigation in the Sekhon et al. framework facilitated the investigation of defined healthcare acceptability constructs from diverse perspectives and data collection methods. Examining multiple viewpoints can provide lessons about the extent to which respondents are good judges of their own and others’ acceptance and capabilities. The joint display enabled cross-source comparison to highlight convergence or divergence in perspective and experience, rather than triangulation.

Intervention coherence was well understood by participants and providers. This knowledge likely was imparted by study staff and indicates a strong ability to convey its mechanism and objectives. Both groups believed providing mental and emotional support was an objective of the intervention. Affective attitude was positive, however, while participants reported nearly universal satisfaction, providers held more diverse viewpoints. Providers indicated that they would be less willing to refer patients versus family members for the interventions. We discovered that this was due to the perceived higher levels of education and support available to healthcare providers’ relatives. These findings indicate that while providers report a positive attitude towards the intervention, that did not translate into willingness to refer patients in the future, despite seeing participants successfully engage with interventions.

Participants were more likely to perceive the interventions as effective than healthcare providers. Provider perceptions of effectiveness were informed by perceived intervention objectives, belief in remote monitoring effectiveness and appropriateness, and perceptions of participant experience. Multiple SESAL physicians perceived reduction in unnecessary return patient visits. Although data were not collected to confirm this, similar findings have been observed in other remote patient monitoring studies, where reduced return visits are frequently cited as a positive outcome.(28)

Providers anticipated remote monitoring would substantially burden patients. Most study participants described a minimal burden and valued the education and support provided. The mismatch between provider perception and participant experience could be due an under-estimation of participant capacity by providers and/or participants underreporting challenges.

Providers identified possible barriers participants may have experienced in their return for evaluation: burden (cost, transportation, accompaniment, time) and emotional responses to interacting with the healthcare system (fear of hospitalization, dying, mistreatment, or disbelief in warning sign severity). While similar participant information is unavailable, of the 52 participants who reported seeking care after a referral, 24 (46%) did not do so. The reasons are unclear, but could be due to social desirability bias or barriers experienced while seeking care. The appropriateness of relying on participants to return for additional care without providing additional support or linkage to care is uncertain, given significant barriers to access.

Most participants reported no difficulty with participation, however, the odds of reporting difficulty significantly increased relative to educational attainment. Participants successfully measured and reported pulse oximeter readings, with at least one measurement provided on 97% of calls, an indication of appropriately held perceptions of self-efficacy. While providers expressed doubts about participant self-efficacy first, many became convinced that participants were capable of self-administering pulse oximetry. However, positivity during in-depth interviews does not negate the anonymous CASI data, where SESAL and study providers indicated greater willingness to refer family than patients for intervention participation, due to doubts about capacity to perform and understand intervention procedures.

While appropriateness was not queried directly with participants, positive affective attitude and perceived efficacy provide some evidence that they believed the intervention was the right fit for the problem. The results of this study raise concerns about the appropriateness of referring participants for additional care without facilitating that process, given low rates of successful return for care. Additionally, two providers provided a systems perspective, raising concerns about whether this intervention would be feasible and a suitable investment to provide remote care for COVID-19 patients, given other, non-COVID-19, health priorities.

Remote patient monitoring has been shown to be a safe and effective way for stable patients at high-risk for adverse outcomes to receive care, with or without the addition of self-administered technology, as well as reducing health facility burden, risk of nosocomial infection, and cost.(29,30) Staffing such an intervention is challenging because infection rates and staffing needs increase concurrently, however, with an emergency preparedness plan and training, healthcare staff were willing and believed themselves capable of implementing these strategies.(31) Previous concerns included lack of access to technology, unreliable mobile and electricity networks, and low health and technology literacy, which may be exacerbated among marginalized groups. (29,32–37) High participation rates demonstrate the penetration of mobile phone technology and the ability of participants to access a phone, even if they do not own one. While concerns about equity in healthcare technology are vital and warranted, assumptions about the capacity of people with lower educational attainment to learn and use new technology should be based in experiences with the target population, rather than assumptions or perceptions, even those of experienced healthcare providers.

With those considerations in mind, intervention adaptation will be crucial if these interventions are adopted in the future. Avenues for adaptation are expansive and might include implementation during future health emergencies to monitor high-risk, stable patients, reduce onward transmission, and conserve health facility resources or adaptation to provide short term remote monitoring of patient post-acute health event, such as monitoring blood glucose after a diabetes diagnosis, including with internet-enabled devices.(38–40) Conceptualizing this approach broadly expands its utility and potential applications, including adapting methods of patient contact and warning sign identification based on the context, patient population, and disease manifestation. Future implementers and researchers adapting this remote monitoring and warning sign identification intervention could focus on: (1) selecting a disease, infectious or chronic, with warning signs detectable through self-administered technology; (2) identifying a medically stable yet high-risk population; (3) collaborating with the target group to ensure appropriate communication and technology; (4) developing an educational approach; (5) addressing provider concerns about patient ability to use self-monitoring tools; and (6) tracking provider referral rates to prevent doubts about self-efficacy from limiting participation. Including mental and emotional health support may also enhance patient and provider satisfaction.

The rigorous methodology of this investigation allows the presentation of credible and transferable results, although further investigation would be needed to understand perspectives and capacities a different target population. This study has limitations. The lack of qualitative investigation with participants limits our understand of the rationale behind their perceptions of the interventions. Additionally, because study staff administered the discharge questionnaires, including questions about acceptability, social desirability bias could positively skew participant responses. Limits of the CASI include the restriction of enrollment to SESAL staff enrollment to those present on data collection days, which excludes SESAL staff not working on those days or during that shift, and those who no longer worked at the triage center when data collection was carried out. In-depth interview limitations include that study personnel conducted the interviews, which could positively bias in-depth interview responses, although they were conducted by senior study staff, rather than staff who previously provided patient care. Participant engagement beyond a ten-day period, or beyond the acute phase of a health concern, is unknown, and could decrease due to a reduced sense of urgency. It is possible that high participation levels were mediated by the isolation mandate for COVID-19 positive patients, where people were eager to participate, talk to study staff, and report symptoms because they were socially isolated during their infection. More broadly, within implementation science, a recent review of acceptability assessments found they often lack standardized approaches and rigorous methodologies. While this assessment of acceptability does align with the best practices identified in this review, it does not rely on validated scales to calculate acceptability metrics.(41) Finally, due to their extensive use, the evidence base around the limitations of fingertip pulse oximeters was expanded upon during the COVID-19 pandemic, leading to an FDA investigation. (42–44) Pulse oximeters were found to measure Sp0_2_ 1.1% lower among self-identified Black patients and 1.2% lower self-identified non-Black Hispanics, findings that are likely applicable to Hondurans and study participants, 97% of whom self-identified as Mestizo or Mestizo and an additional ethnicity, most frequently Lenca or Maya, during enrollment. While the relatively high referral threshold of 94% Sp0_2_ may have avoided under-referrals, better understanding the impact of skin color on pulse oximeter precision, and adjusting warning and referral levels appropriately is essential prior to their use in a population that includes members with darker skin, to avoid delays in identification of hypoxia and referral to appropriate care.(45)

In summary, while the intervention was designed to mitigate COVID-19-associated morbidity and mortality, widespread adoption is no longer needed due to the pronounced decrease in severe outcomes. Nonetheless, remote patient monitoring and self-administered technology are powerful tools that can be leveraged to provide remote monitoring of acute conditions, such as when systems are overwhelmed during future epidemics or post-acute health events, whether due to chronic or infectious diseases. Our findings provide evidence for how these interventions may be implemented in a lower resource setting during a pandemic, and how patients and implementors perceive these interventions. Given that successful implementation of healthcare interventions can be grounded in how they are perceived, generating rigorous data to understand and dissect these perceptions is a critical building block in preparing for the next epidemic or pandemic.

## Data Availability

Data are available upon reasonable request, as they are owned by the Secretariat of Health of Honduras.

## 5 Acknowledgements

Our sincere gratitude to the Honduras Secretaría de Salud, including the Health Networks and Surveillance divisions, staff of COVID-19 screening centers and the Social Security Hospital, and study staff in Honduras, all of whom worked tirelessly to provide high-quality care. Thank you to the participants who kindly shared their time and perspectives. Our appreciation to Roche Diagnostics Ltd. for donating rapid COVID-19 antigen tests through an Investigator Initiated Study.

## 6 Supplementary materials

### S1 COVID-19 Risk factors – as defined by the U.S. CDC as of March 2021(20)

- Age ≥ 60 years
- Cancer
- Cerebrovascular disease (stroke or transient ischemic attack)
- Chronic renal disease
- Chronic pulmonary disease (except asthma)
- Chronic kidney disease
- Diabetes mellitus, type I or II
- Heart disease (cardiac insufficiency, myocardiopathies, coronary artery disease)
- Mental illness limited to depression, bipolar, or schizophrenia
- Hypertension
- Obesity (BMI ≥ 30 kg/m^2^)
- Pregnancy or recent pregnancy (< 1 month)
- Tobacco use, current or previous
- Tuberculosis

### S2 Trial participation refusers vs. participants

S2 - Trial participation refusers vs. participants
|  | Refused | Enrolled | OR [CI] | p value |
| --- | --- | --- | --- | --- |
| **Refused vs Enrolled** | 1.0% (n=19) | 98.9% (n=1,821) |  |  |
| **Study intervention** |  |  |  |  |
| Monitoring only | 63.2% (n=12) | 50.7% (n=924) | Ref. |  |
| Pulse oximetry | 36.8% (n=7) | 49.3% (n=897) | 0.60 [0.23 - 1.52] | 0.287 |
| **Gender** |  |  |  |  |
| Female | 36.8% (n=7) | 60.8% (n=1,106) | Ref. |  |
| Male | 58.9% (n=11) | 39.1% (n=711) | 2.43 [CI 0.94 - 6.30] | 0.068 |
| Non-binary | 0% (n=0) | 0.05% (n=1) | Omitted |  |
| Other | 5.3% (n=1) | 0.05% (n=1) | 159 [9.0 – 2808.5] | 0.001 |
| **Age (median [IQR])** | 59.4 [6.0] | 61.6 [10.3] | 0.98 [CI 0.93 - 1.02] | 0.342 |
| **Health status at enrollment** |  |  |  |  |
| Temperature (mean [SD]) | 36.4 [0.36] | 36.5 [0.46] | 0.46 [0.200 - 1.09] | 0.078 |
| SpO2 ≤ 94% (mean [SD]) | 96.9 [1.27] | 96.7 [1.27] | 1.16 [0.81 - 1.68] | 0.418 |
| Heart rate (mean [SD]) | 85.3 [10.4] | 85.2 [12.8] | 1.00 [0.97 - 1.04] | 0.984 |
| Respiratory rate (mean [SD]) | 17.6 [1.7] | 18.1 [1.7] | 0.85 [0.67 - 1.08] | 0.181 |

### S3 CASI respondent characteristics

S3 - CASI respondent characteristics
| Gender |  | Female |  |  | Male |  |  | Total |
| --- | --- | --- | --- | --- | --- | --- | --- | --- |
| Role | Age | 20- 29 | 30 – 39 | 40 – 59 | 20- 29 | 30 – 39 | 40 – 59 |  |
| SESAL – Physician administrator |  | 0 | 1 | 0 | 0 | 2 | 0 | 3 |
| SESAL – Attending physician |  | 3 | 6 | 1 | 0 | 5 | 1 | 16 |
| Study – Medical interviewer |  | 2 | 4 | 0 | 0 | 2 | 0 | 8 |
| Study – Monitoring nurse |  | 1 | 4 | 1 | 1 | 0 | 0 | 7 |
| **Total** |  | **6** | **15** | **2** | **1** | **9** | **1** | **34** |

### S4 In-depth interview respondent characteristics

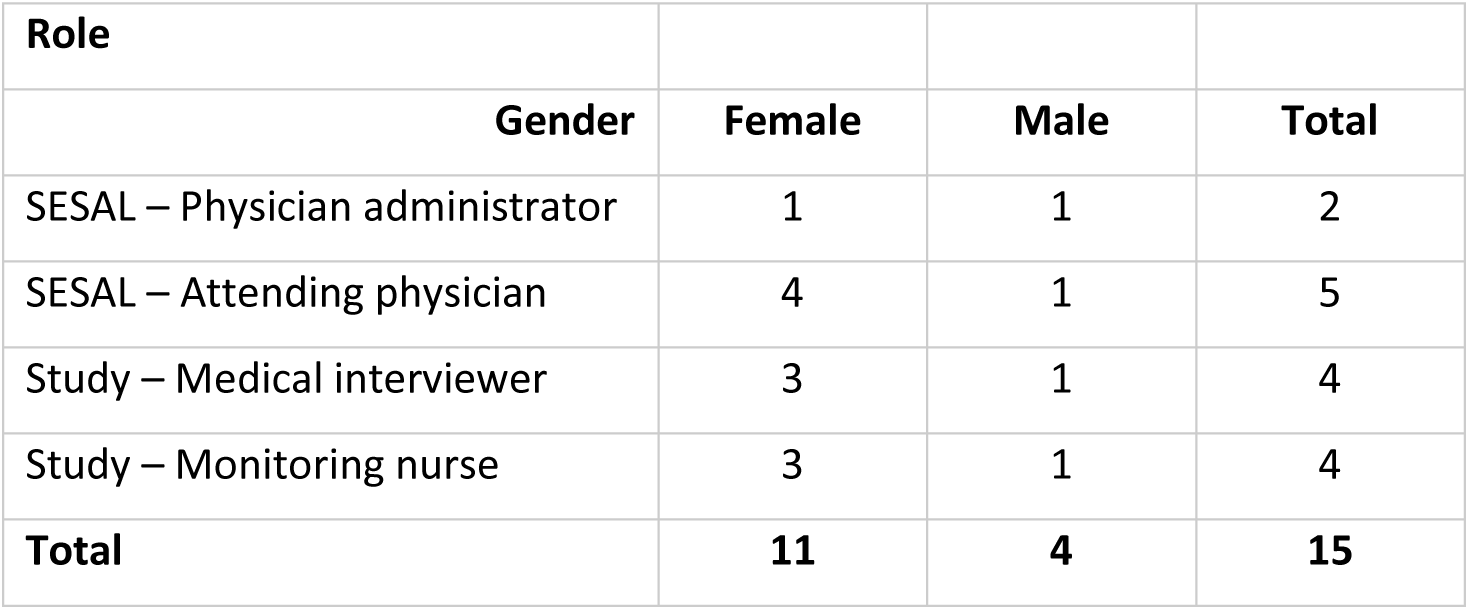

## References

1. Nuzzo JB, Meyer D, Snyder M, Ravi SJ, Lapascu A, Souleles J, et al. What makes health systems resilient against infectious disease outbreaks and natural hazards? Results from a scoping review. BMC Public Health. 2019 Oct 17;19(1):1310.

2. Zhao L, Jin Y, Zhou L, Yang P, Qian Y, Huang X, et al. Evaluation of health system resilience in 60 countries based on their responses to COVID-19. Front Public Health [Internet]. 2023 Jan 9 [cited 2024 Jun 19];10. Available from: https://www.frontiersin.org/journals/public-health/articles/10.3389/fpubh.2022.1081068/full

3. Anaya-Covarrubias JY, Pizuorno A, Mirazo S, Torres-Flores J, Du Pont G, Lamoyi E, et al. COVID-19 in Latin America and the caribbean region: Symptoms and morbidities in the epidemiology of infection. Curr Opin Pharmacol. 2022 Apr;63:102203.

4. World Health Organization. Home care for patients with suspected or confirmed COVID-19 and management of their contacts [Internet]. 2020 [cited 2022 Nov 11]. Available from: https://www.who.int/publications-detail-redirect/home-care-for-patients-with-suspected-novel-coronavirus-(ncov)-infection-presenting-with-mild-symptoms-and-management-of-contacts

5. Nilles EJ. Implementation and Evaluation of Home-based Care and Hand Hygiene Interventions in Honduras [Internet]. clinicaltrials.gov; 2022 Nov [cited 2023 Dec 31]. Report No.: NCT04886414. Available from: https://clinicaltrials.gov/study/NCT04886414

6. Secretaria de Salud de Honduras (SESAL). Guía Práctica Para El Manejo Domiciliar De Pacientes Sospechosos o Confirmados por COVID-19. Tegucigalpa, Honduras; 2022 Mar p. 92. Report No.: Rev. 01-22.

7. Salud S de. gob.mx. [cited 2023 Nov 25]. Avanza el uso de la Telesalud o Telemedicina en México. Available from: http://www.gob.mx/salud/prensa/avanza-el-uso-de-la-telesalud-o-telemedicina-en-mexico

8. Carvalho CRR, Scudeller PG, Rabello G, Gutierrez MA, Jatene FB. Use of telemedicine to combat the COVID-19 pandemic in Brazil. Clinics. 2020 Aug 3;75:e2217.

9. El auge de la telemedicina en medio de la COVID-19 [Internet]. Ideas que Cuentan. 2021 [cited 2023 Nov 25]. Available from: https://blogs.iadb.org/ideas-que-cuentan/es/el-auge-de-la-telemedicina-en-medio-de-la-covid-19/

10. Moya Díaz GM. Ethical Telemedicine for Honduras in times of COVID-19. Rev Cienc Forenses Honduras. 2020 Dec 30;6(2):38–45.

11. Alipour J, Hayavi-Haghighi MH. Opportunities and Challenges of Telehealth in Disease Management during COVID-19 Pandemic: A Scoping Review. Appl Clin Inform. 2021 Aug;12(4):864–76.

12. Rijal S, Poudel B. Is home pulse oximeter monitoring for COVID-19 feasible in low-income and low-middle-income countries? BMJ Health Care Inform. 2021 Aug 1;28(1):e100465.

13. Baker K, Ward C, Maurel A, Cola M, Smith H, Habte T, et al. Usability and acceptability of a multimodal respiratory rate and pulse oximeter device in case management of children with symptoms of pneumonia: A cross-sectional study in Ethiopia. Acta Paediatrica. 2020 Dec 1;110.

14. Eccles MP, Mittman BS. Welcome to Implementation Science. Implementation Science. 2006 Feb 22;1(1):1.

15. Bauer MS, Damschroder L, Hagedorn H, Smith J, Kilbourne AM. An introduction to implementation science for the non-specialist. BMC Psychol. 2015 Sep 16;3(1):32.

16. Proctor E, Silmere H, Raghavan R, Hovmand P, Aarons G, Bunger A, et al. Outcomes for Implementation Research: Conceptual Distinctions, Measurement Challenges, and Research Agenda. Adm Policy Ment Health. 2011;38(2):65–76.

17. Sekhon M, Cartwright M, Francis JJ. Acceptability of healthcare interventions: an overview of reviews and development of a theoretical framework. BMC Health Services Research. 2017 Jan 26;17(1):88.

18. Creswell JW, Clark VLP. Designing and Conducting Mixed Methods Research. Second edition. Los Angeles: SAGE Publications, Inc; 2010. 488 p.

19. Fetters MD, Freshwater D. The 1 + 1 = 3 Integration Challenge. Journal of Mixed Methods Research. 2015 Apr 1;9(2):115–7.

20. The Centers for Disease Control and Prevention. Centers for Disease Control and Prevention. 2023 [cited 2023 Jul 17]. People with Certain Medical Conditions. Available from: https://www.cdc.gov/coronavirus/2019-ncov/need-extra-precautions/people-with-medical-conditions.html

21. KoboToolbox [Internet]. [cited 2022 Sep 29]. KoboToolbox | Data Collection Tools for Challenging Environments. Available from: https://www.kobotoolbox.org/

22. StataCorp. Stata Statistical Software: Release 17. College Station, TX: StataCorp LLC; 2021.

23. R Core Team. R: A language and environment for statistical computing. [Internet]. Vienna, Austria: R Foundation for Statistical Computing; 2022. Available from: https://www.R-project.org/

24. Richards K, Hemphill M. A Practical Guide to Collaborative Qualitative Data Analysis. Journal of Teaching in Physical Education. 2017 Oct 16;37:1–20.

25. Dedoose [Internet]. Los Angeles, CA: SocioCultural Research Consultants, LLC; Available from: www.dedoose.com

26. Microsoft Excel Spreadsheet Software | Microsoft 365 [Internet]. [cited 2022 Dec 31]. Available from: https://www.microsoft.com/en-us/microsoft-365/excel

27. Guetterman TC, Fetters MD, Creswell JW. Integrating Quantitative and Qualitative Results in Health Science Mixed Methods Research Through Joint Displays. Ann Fam Med. 2015 Nov;13(6):554–61.

28. Vudathaneni VKP, Lanke RB, Mudaliyar MC, Movva KV, Mounika Kalluri L, Boyapati R. The Impact of Telemedicine and Remote Patient Monitoring on Healthcare Delivery: A Comprehensive Evaluation. Cureus. 16(3):e55534.

29. Camacho-Leon G, Faytong-Haro M, Carrera K, Molero M, Melean F, Reyes Y, et al. A Narrative Review of Telemedicine in Latin America during the COVID-19 Pandemic. Healthcare (Basel). 2022 Jul 22;10(8):1361.

30. Mahmoud K, Jaramillo C, Barteit S. Telemedicine in Low- and Middle-Income Countries During the COVID-19 Pandemic: A Scoping Review. Front Public Health. 2022 Jun 22;10:914423.

31. Task shifting healthcare services in the post-COVID world: A scoping review | PLOS Global Public Health [Internet]. [cited 2024 May 15]. Available from: https://journals.plos.org/globalpublichealth/article?id=10.1371/journal.pgph.0001712

32. Vindrola-Padros C, Sidhu MS, Georghiou T, Sherlaw-Johnson C, Singh KE, Tomini SM, et al. The implementation of remote home monitoring models during the COVID-19 pandemic in England. EClinicalMedicine. 2021 Apr;34:100799.

33. Gaeta T, Chiricolo G, Mendoza C, Vaccari N, Melville L, Melniker L, et al. Impact of a Novel Telehealth Follow-Up Protocol for At-Risk Emergency Department Patients Discharged With Presumptive or Confirmed COVID-19. Annals of Emergency Medicine. 2020 Oct 1;76(4):S49.

34. Diseases TLI. Unmet need for COVID-19 therapies in community settings. The Lancet Infectious Diseases. 2021 Nov 1;21(11):1471.

35. Agarwal P, Mukerji G, Laur C, Chandra S, Pimlott N, Heisey R, et al. Adoption, feasibility and safety of a family medicine–led remote monitoring program for patients with COVID-19: a descriptive study. CMAJ Open. 2021 Mar 30;9(2):E324–30.

36. Maghrabi F, Bazaz R, Wilson E, O’Reilly S, Calisti G, Richardson R, et al. S57 The development and implementation of a virtual discharge ward for patients with COVID-19 pneumonia: data on the first 300 patients. Thorax. 2021 Feb 1;76(Suppl 1):A35–6.

37. Arora P, Mehta D, Ha J. Impact of telehealth on health care resource utilization during the COVID-19 pandemic. Journal of Comparative Effectiveness Research. 2022 Apr;11(5):301–9.

38. Alboksmaty A, Beaney T, Elkin S, Clarke JM, Darzi A, Aylin P, et al. Effectiveness and safety of pulse oximetry in remote patient monitoring of patients with COVID-19: a systematic review. The Lancet Digital Health. 2022 Apr 1;4(4):e279–89.

39. DeRogatis MJ, Pellegrino AN, Wang N, Higgins M, Dubin J, Issack P, et al. Enhancing recovery and reducing readmissions: The impact of remote monitoring on acute postoperative care in outpatient total joint arthroplasty. Journal of Orthopaedics. 2024 Dec 1;58:111–6.

40. Salehi S, Olyaeemanesh A, Mobinizadeh M, Nasli-Esfahani E, Riazi H. Assessment of remote patient monitoring (RPM) systems for patients with type 2 diabetes: a systematic review and meta-analysis. J Diabetes Metab Disord. 2020 Jun 1;19(1):115–27.

41. Ortblad KF, Sekhon M, Wang L, Roth S, van der Straten A, Simoni JM, et al. Acceptability Assessment in HIV Prevention and Treatment Intervention and Service Delivery Research: A Systematic Review and Qualitative Analysis. AIDS Behav. 2023 Feb;27(2):600–17.

42. Food and Drug Administration. FDA. 2022 [cited 2022 Oct 23]. November 1, 2022: Anesthesiology and Respiratory Therapy Devices Panel of the Medical Devices Advisory Committee Meeting Announcement - 11/01/2022. Available from: https://www.fda.gov/advisory-committees/advisory-committee-calendar/november-1-2022-anesthesiology-and-respiratory-therapy-devices-panel-medical-devices-advisory

43. Health C for D and R. Pulse Oximeter Accuracy and Limitations: FDA Safety Communication. FDA [Internet]. 2022 Sep 14 [cited 2022 Oct 23]; Available from: https://www.fda.gov/medical-devices/safety-communications/pulse-oximeter-accuracy-and-limitations-fda-safety-communication

44. McFarling UL. Pulse oximeters’ inaccuracies in darker-skinned people require urgent action, AGs tell FDA [Internet]. STAT. 2023 [cited 2023 Nov 8]. Available from: https://www.statnews.com/2023/11/07/pulse-oximeters-attorneys-general-urge-fda-action/

45. Fawzy A, Wu TD, Wang K, Robinson ML, Farha J, Bradke A, et al. Racial and Ethnic Discrepancy in Pulse Oximetry and Delayed Identification of Treatment Eligibility Among Patients With COVID-19. JAMA Internal Medicine. 2022 Jul 1;182(7):730–8.

